# Tenecteplase versus alteplase for patients with minor acute ischemic stroke: an analysis of the ORIGINAL clinical trial

**DOI:** 10.64898/2026.03.17.26348663

**Authors:** Shuhong Xu, Hongguo Dai, Guozhi Lu, Weiwei Wang, Fengyuan Che, Yu Geng, Xiaolong Bao, Shijia Yan, Shuya Li, Yongjun Wang

**Affiliations:** Department of Neurology and Department of Clinical Trial Center, Beijing Tiantan Hospital, Capital Medical University, Beijing, China; China National Clinical Research Center for Neurological Diseases, Beijing, China; Linfen Central Hospital, Linfen, China; Hexigten Banner Mongolian Traditional Chinese Medicine Hospital, Chifeng, China; Xianyang Hospital of Yan’an University, Xianyang, China; Linyi People’s Hospital, Linyi, China; Zhejiang Provincial People’s Hospital, Hangzhou, China; Boehringer Ingelheim, Shanghai, China

**Keywords:** acute ischemic stroke, alteplase, intravenous thrombolysis, minor stroke, tenecteplase

## Abstract

**Background:** Stroke guidelines recommend intravenous thrombolysis (IVT) within 4.5 hours of symptom onset for patients with minor acute ischemic stroke (AIS) but disabling symptoms. However, such patients are often overlooked for treatment, increasing their risk of stroke-related disability. Tenecteplase is endorsed as an alternative to alteplase for IVT in patients with AIS. More evidence is required regarding its efficacy and safety in the minor stroke population.

**Methods:** This post hoc analysis of the ORIGINAL randomized clinical trial aimed to evaluate the efficacy and safety of tenecteplase versus alteplase in the patient subgroup with minor (National Institutes of Health Stroke Scale [NIHSS] ≤5) disabling stroke. Primary outcome was the proportion of patients with a modified Rankin Scale (mRS) score of 0 or 1 at Day 90.

**Results:** Data were analyzed for 299 patients treated with tenecteplase 0.25 mg/kg and 297 patients treated with alteplase 0.9 mg/kg. At Day 90, 86.3% of tenecteplase recipients and 82.8% of alteplase recipients achieved a mRS score of 0 or 1 (risk ratio=1.04 [95% confidence interval 0.971–1.114]; non-significant). No heterogeneity of treatment effect was observed across predefined subgroups according to baseline NIHSS score, time to drug administration, sex, age, presence (yes/no) of atrial fibrillation and diabetes and thrombectomy performed. No statistically significant differences were observed between tenecteplase and alteplase across secondary efficacy and safety outcomes.

**Conclusions:** The comparable efficacy and safety of tenecteplase 0.25 mg/kg and alteplase 0.9 mg/kg in the minor stroke population of the ORIGINAL randomized clinical trial suggests that tenecteplase is a suitable alternative to alteplase in this setting.

**Trial registration:** ClinicalTrials.gov NCT04915729 (ORIGINAL randomized clinical trial; https://clinicaltrials.gov/study/NCT04915729). Submitted 4 June 2021.

## Introduction

More than half of patients with acute ischemic stroke (AIS) present with mild neurological deficits.^1^ Mild symptoms are a common reason for nonuse of intravenous thrombolysis (IVT); however, a substantial proportion of untreated patients are at risk of residual disability.^2−4^ Large-scale analyses have demonstrated that IVT in patients with minor stroke is associated with an increased likelihood of a favorable functional outcome.^1,5−8^ International and local stroke guidelines recommend IVT within 4.5 hours of symptom onset for patients with minor stroke but disabling symptoms.^9−11^

Recombinant tissue plasminogen activator (rtPA; hereafter, alteplase) has been the primary thrombolytic agent for AIS for more than 30 years. Tenecteplase (TNK) is a third-generation tissue plasminogen activator with improved pharmacological properties including greater fibrin specificity, longer half-life and lower plasma clearance, facilitating administration as a single bolus dose.^12^ Meta-analyses of head-to-head randomized controlled trials (RCTs) of tenecteplase versus alteplase in AIS report that tenecteplase is superior for achieving excellent functional outcomes.^13,14^ Tenecteplase is recommended as an alternative to alteplase within 4.5 hours of symptom onset in patients with AIS who are eligible for IV thrombolytics.^15−17^

The substantial size and unmet needs of the minor stoke population underscore the importance of identifying treatment options for this patient group. An individual patient data meta-analysis of randomized trials of thrombolysis with intravenous alteplase was first to support use of IVT for patients with minor disabling stroke.^5^ Subsequent retrospective real-world evidence confirmed that patients with minor non-disabling stroke patients with large vessel occlusion also benefit from alteplase thrombolysis.^18^ The TEMPO-1/2 RCTs explored the safety and efficacy of tenecteplase within 12 hours of symptom onset in patients with minor stroke and confirmed its safety, confirmed the safety of tenecteplase in this population.^19,20^ Currently, evidence regarding the safety and efficacy of tenecteplase in minor stroke within the 4.5-hour post-onset window remains insufficient.

The phase 3 ORIGINAL randomized clinical trial reported comparable efficacy and safety outcomes with tenecteplase and alteplase in Chinese patients with AIS and measurable neurological deficit.^21^ This post hoc analysis of the ORIGINAL clinical trial was undertaken to evaluate the efficacy and safety of tenecteplase versus alteplase in the minor stroke subgroup (National Institutes of Health Stroke Scale [NIHSS] ≤5) with the goal of providing additional evidence to support treatment selection for patients with minor disabling stroke.

## Methods

### Study Design

The ORIGINAL randomized clinical trial (NCT04915729) was a multicenter, active-controlled, parallel-group, randomized, open-label, blinded endpoint trial that assessed the non-inferiority of tenecteplase (Boehringer Ingelheim) to alteplase in Chinese patients with AIS within 4.5 hours of symptom onset. Methods and results have been reported elsewhere.^21^ Briefly, patients were recruited from 55 neurology clinics and stroke centers in China and were eligible if they had AIS with a NIHSS score of 1–25, measurable neurological deficit and were symptomatic for at least 30 minutes without significant improvement. Patients were randomized (1:1) within 4.5 hours of symptom onset to receive intravenous (IV) tenecteplase 0.25 mg/kg as a bolus over 5 to 10 seconds or IV alteplase 0.9 mg/kg with 10% of the dose administered as an initial bolus and the remainder administered immediately after as an infusion over 1 hour.

This post hoc analysis was undertaken to investigate efficacy and safety outcomes in the patient subgroup with a NIHSS score ≤5: either a NIHSS score of <4 plus measurable deficit of at least 1 in the motor function score for arms or legs or a NIHSS score of 4−5 with no other requirement. The primary outcome was the proportion of patients with a modified Rankin Scale (mRS) score of 0 (no symptoms at all) or 1 (no significant disability despite symptoms; able to carry out all usual duties and activities) at Day 90. Secondary efficacy outcomes were major neurological improvement at 24 hours, mRS score of 0–2 at Day 90, change in NIHSS score from baseline to Day 90, distribution of mRS scores at Day 90, and Barthel Index score of ≥95 at Day 90. Secondary safety outcomes were on-treatment symptomatic intracerebral hemorrhage (sICH) per European Cooperative Acute Stroke Study (ECASS) III, 90-day all-cause mortality, and mRS score of 5 or 6 at Day 90.

### Statistical Analysis

Statistical methods for the post hoc analysis are as reported for the ORIGINAL clinical trial.^21^ Briefly, patients’ baseline characteristics are summarized descriptively using mean ± standard deviation or median (range) for continuous variables and number (percentage) for categorical variables. The primary outcome was analyzed using a log binomial regression model adjusted for continuous covariates (baseline NIHSS score, age, and time to drug administration since onset of AIS) and transformed into a risk ratio (RR) along with 95% confidence intervals (CI). The primary outcome was also analyzed in predefined population subgroups: baseline NIHSS score (<4, ≥4), time to drug administration (≤3, >3 hours), sex (male, female), age (≤80, >80 years), atrial fibrillation (yes, no), diabetes (yes, no) and thrombectomy performed (yes, no).

A similar model adopted for analysis of the primary outcome was used to analyze the secondary efficacy functional outcomes of major neurological improvement at 24 hours, mRS score of 0–2 at Day 90, and Barthel Index score of ≥95 at Day 90, with results reported as RR with 2-sided 95% CIs. For change from baseline in NIHSS score at day 90, the mixed model repeated measures approach was employed, with results presented as mean (standard error) for each treatment group and as adjusted mean difference (95% CI) for tenecteplase versus alteplase. For distribution of mRS scores at day 90, an ordinal logistic regression model was used including treatment as main effect and baseline NIHSS score, age, and time to study drug administration as continuous covariates, stated as an odds ratio (95% CI). The proportion of patients with adjudicated sICH per the ECASS III definition and all-cause mortality within 90 days while receiving treatment was analyzed using the Suissa-Shuster test and χ2 test, respectively. The proportion of patients with a mRS score of 5 or 6 on day 90 was analyzed using the log-binomial regression model adjusted for continuous covariates.

A two-sided p<0.05 was considered statistically significant. As potential exists for a type 1 error due to multiple statistical tests without adjustment of p values, all analyses should be considered exploratory. As per the main ORIGINAL clinical trial, all statistical analyses were performed using SAS version 9.4 (SAS Institute, Cary, NC, USA).

## Results

### Characteristics of Minor Stroke Participants

Of 1465 patients in the full analysis set of the ORIGINAL randomized clinical trial, 596 (tenecteplase n=299, alteplase n=297) had NIHSS ≤5 and were included in the post hoc analysis. Baseline characteristics were balanced between the tenecteplase and alteplase treatment groups (Table 1). Median age was 65 (range: 58-72) years and median NIHSS score was 4 (range 1−5) in both groups. Comorbidities were common, especially hypertension which was present in 70.6% and 76.1% of patients treated with tenecteplase and alteplase, respectively.

**Table 1.** Baseline characteristics of ORIGINAL clinical trial participants with minor stroke (NIHSS ≤5) treated with tenecteplase or alteplase.

| Baseline characteristic | Tenecteplase (n = 299) | Alteplase (n = 297) |
| --- | --- | --- |
| Age (years), median (IQR) | 65 (58.0–72.0) | 65 (58.0–72.0) |
| Sex, n (%) |  |  |
| Male | 223 (74.6) | 206 (69.4) |
| Female | 76 (25.4) | 91 (30.6) |
| Comorbidities, n (%) |  |  |
| Hypertension | 211 (70.6) | 226 (76.1) |
| Diabetes | 85 (28.4) | 86 (29.0) |
| Coronary artery disease | 52 (17.4) | 53 (17.8) |
| Atrial fibrillation | 32 (10.7) | 25 (8.4) |
| NIHSS, median (range) | 4.0 (1–5) | 4.0 (1–5) |
| Prior stroke in the last 3 months | 4 (1.3) | 7 (2.4) |
| Time to study drug administration, h |  |  |
| $\leq 3$ | 153 (51.2) | 142 (47.8) |
| $> 3$ | 146 (48.9) | 155 (52.3) |
| IQR, interquartile range; NIHSS, National Institutes of Health Stroke Scale. |  |  |

### Outcomes

At Day 90, 86.3% (258/299) of patients receiving tenecteplase and 82.8% (246/297) of patients receiving alteplase achieved the primary outcome (mRS score of 0–1; RR=1.04 [95% CI 0.971–1.114]). There was no significant difference between the tenecteplase and alteplase groups (Figure 1).

**Figure 1.**
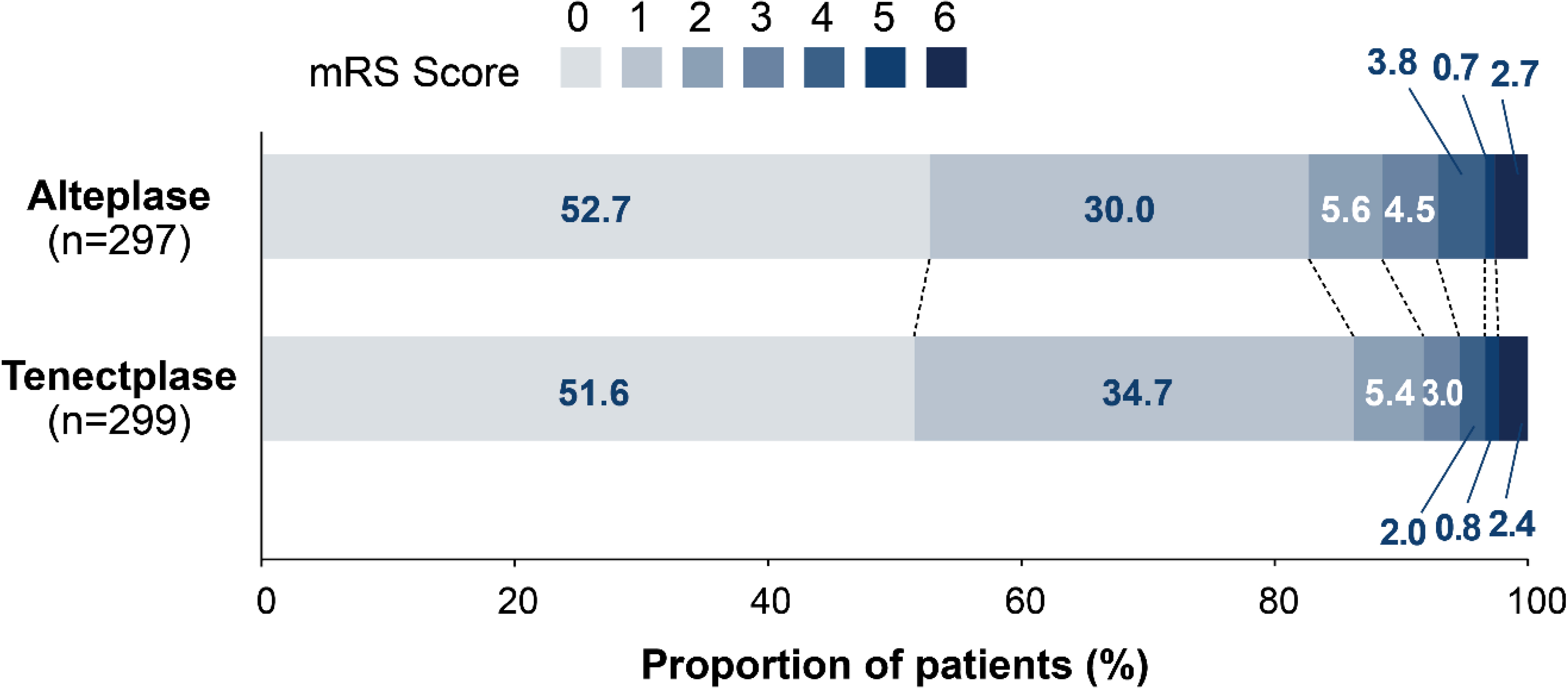
Primary efficacy outcome: distribution of modified Rankin Scale (mRS) scores at Day 90 in ORIGINAL study participants with minor stroke (NIHSS ≤5) treated with tenecteplase or alteplase. NIHSS, National Institutes of Health Stroke Scale.

By predefined subgroup, including baseline NIHSS score, time to drug administration, sex, age, and presence (yes, no) of atrial fibrillation and diabetes, point estimates for the primary efficacy outcome were greater than 1, favoring tenecteplase (Figure 2).

**Figure 2.**
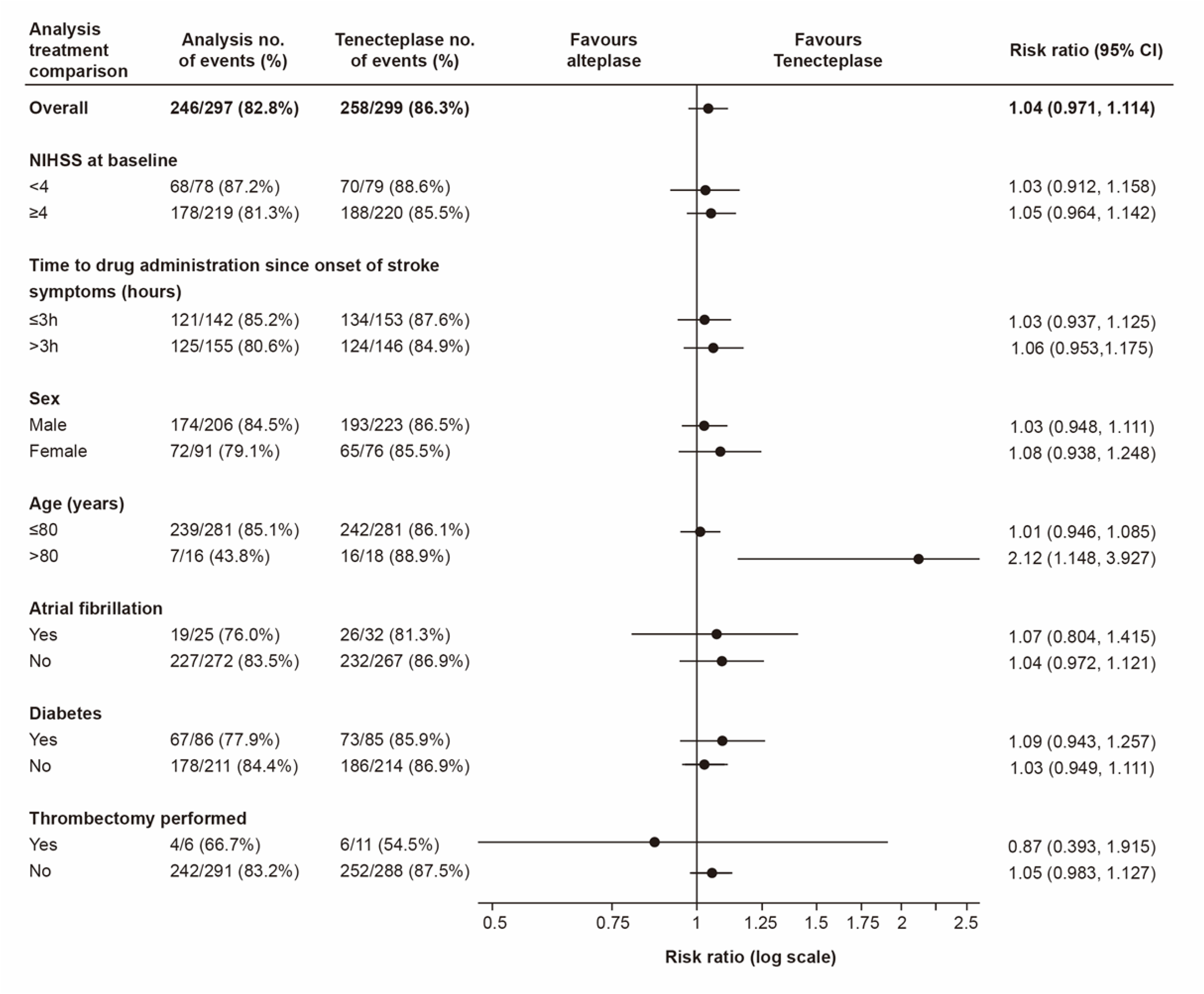
Forest plot of risk ratios for modified Rankin Scale score of 0 or 1 on Day 90 (primary outcome measure) stratified by predefined subgroups among ORIGINAL study participants with minor stroke (NIHSS ≤5) treated with tenecteplase or alteplase. The log binomial regression model was adjusted for continuous covariates (baseline NIHSS score, age, and time to drug administration since onset of AIS) and transformed into a risk ratio along with 95% CIs. AIS, acute ischemic stroke; CI, confidence interval; NIHSS, National Institutes of Health Stroke Scale.

Results for secondary efficacy outcomes were similar between tenecteplase and alteplase with no significant differences between treatment groups (Table 2). Risk ratios for major neurological improvement at 24 h, mRS score 0–2 on Day 90, and Barthel index score of at least 95 on Day 90 ranged from 1.03 to 1.13, favoring tenecteplase. The odds ratio (95% CI) of 1.04 (0.873−1.229) for distribution of mRS score on Day 90 favored tenecteplase. The adjusted mean difference (95% CI) of -0.28 (-1.45−0.88) for change in NIHSS score from baseline to Day 90 favored tenecteplase.

**Table 2.** Secondary efficacy outcomes in ORIGINAL clinical trial participants with minor stroke (NIHSS ≤5) treated with tenecteplase or alteplase.

| Variable | Tenecteplase | Alteplase | Risk ratio* (95% CI) |
| --- | --- | --- | --- |
| Major neurological improvement at 24 h, n/N (%) | 115/291 (39.5) | 103/294 (35.0) | 1.13 (0.914–1.390) |
| mRS score 0–2 on Day 90, n/N (%) | 274/299 (91.6) | 262/297 (88.2) | 1.04 (0.983–1.093) |
| Change in NIHSS score from baseline to Day 90, mean (SE) | -2.34 (0.41) | -1.84 (0.44) | -0.28<sup>†</sup> (-1.45–0.88) |
| Distribution of mRS score on Day 90 | – | – | 1.04<sup>‡</sup> (0.873–1.229) |
| Barthel index score of $\geq 95$ on Day 90, n/N (%) | 250/286 (87.4) | 245/290 (84.5) | 1.03 (0.965–1.099) |

\* Unless otherwise indicated.
<sup>†</sup> Adjusted mean difference (95% CI).
<sup>‡</sup> Odds ratio (95% CI).
CI, confidence interval; mRS, modified Rankin scale; NIHSS, National Institutes of Health Stroke Scale; SE, standard error.

Safety outcomes were similar between tenecteplase and alteplase groups (Table 3). sICH occurred in 5 patients (1.7%) receiving tenecteplase and 5 patients (1.7%) receiving alteplase (RR=1.00 [95% CI 0.276–3.648]). The 90-day mortality rate was 2.3% with tenecteplase and 2.7% with alteplase (RR=0.88 [95% CI 0.322–2.390]). Ten patients in each treatment group had a mRS score of 5 or 6 at Day 90 (3.3% vs 3.4%, RR=0.94 [95% CI 0.399–2.233]).

**Table 3.**
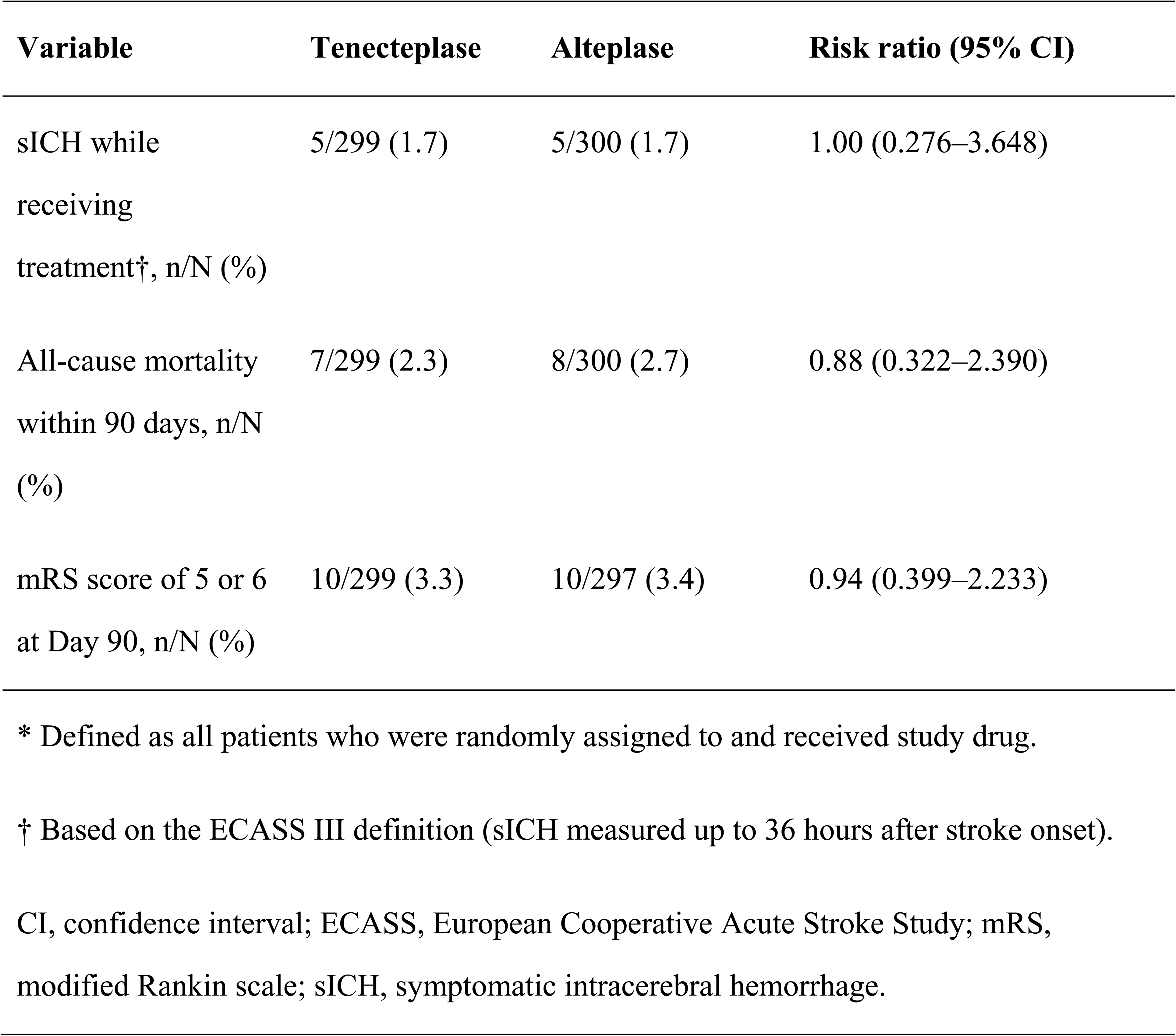
Safety outcomes in ORIGINAL clinical trial participants with minor stroke treated with tenecteplase or alteplase: safety set*.

| Variable | Tenecteplase | Alteplase | Risk ratio (95% CI) |
| --- | --- | --- | --- |
| sICH while receiving treatment†, n/N (%) | 5/299 (1.7) | 5/300 (1.7) | 1.00 (0.276–3.648) |
| All-cause mortality within 90 days, n/N (%) | 7/299 (2.3) | 8/300 (2.7) | 0.88 (0.322–2.390) |
| mRS score of 5 or 6 at Day 90, n/N (%) | 10/299 (3.3) | 10/297 (3.4) | 0.94 (0.399–2.233) |

\* Defined as all patients who were randomly assigned to and received study drug.
† Based on the ECASS III definition (sICH measured up to 36 hours after stroke onset).
CI, confidence interval; ECASS, European Cooperative Acute Stroke Study; mRS, modified Rankin scale; sICH, symptomatic intracerebral hemorrhage.

## Discussion

This post hoc analysis of patients with minor disabling AIS enrolled in the ORIGINAL clinical trial indicated that tenecteplase 0.25 mg/kg and alteplase 0.9 mg/kg were comparable with regard to the proportion of patients with excellent functional outcome (mRS score of 0–1) at 90 days. No statistically significant differences were observed between tenecteplase and alteplase across any secondary efficacy or safety outcomes.

The comparability of tenecteplase and alteplase across all efficacy and safety outcomes in the minor stroke subgroup was consistent with results reported in the overall ORIGINAL population. However, compatible with the association between lower baseline NIHSS scores and better stroke outcomes, rates of excellent functional outcome were higher (86.3% with tenecteplase and 82.8% with alteplase versus 72.7% and 70.3%, respectively, in the overall ORIGINAL population), and rates of 90-day mortality were lower (2.3% with tenecteplase and 2.7% with alteplase versus 4.6% and 5.8%, respectively, in the overall ORIGINAL population), in the minor stroke population. The slightly larger absolute risk difference (3.5% vs 2.4%) in the primary outcome in the minor stroke subgroup compared with the overall ORIGINAL population suggests the possibility of greater benefit with tenecteplase over alteplase in this clinical group, without any increase in mortality risk.

The results of this post hoc analysis align with existing published literature. Similar to our findings, a secondary analysis of patients with minor stroke from the AcT trial reported no significant differences in efficacy and safety outcomes between tenecteplase 0.25 mg/kg and alteplase 0.9 mg/kg. Of 378 enrolled patients with NIHSS ≤5, the primary outcome (mRS score 0−1 at 90 to 120 days) was recorded in 51.8% of patients in the tenecteplase group and 47.5% in the alteplase group (adjusted RR=1.14 [95% CI 0.92−1.40]). Rates of sICH (2.9% vs 3.3%; unadjusted RR=0.79 [95% CI 0.24−2.54]) and death within 90 days (5.5% vs 11%; adjusted HR=0.99 [95% CI 0.96−1.02]) were also comparable between treatment groups.^22^ In both the ORIGINAL and AcT post hoc analyses, results for functional outcomes and mortality at 90 days were numerically in favor of tenecteplase. Interestingly, rates of excellent functional outcome with tenecteplase and alteplase in the ORIGINAL minor stroke population were 2.3-fold higher than those recorded in the AcT minor stroke population. The reasons for this large differential are unclear but may reflect differences in patient populations. The ORIGINAL trial defined minor stroke as NIHSS ≤5 and applied an additional operational definition for those with NIHSS <4, i.e. a measurable deficit in motor function score for the arms or legs of ≥1. In AcT, it was assumed that minor stroke patients had disabling deficits at presentation as this is a requirement to be eligible for thrombolysis as per the Canadian stroke guidelines, but the level of disability was not explicitly defined.

Rates of death within 90 days with tenecteplase and alteplase in the ORIGINAL minor stroke population were 2.4-fold and 4.1-fold lower, respectively, than in the AcT minor stroke population, whereas sICH rates were similar in both analyses, suggesting that causes of death in AcT could be for reasons beyond IVT.

The results of our post hoc analysis supplement real-world evidence documenting the efficacy and safety of tenecteplase and alteplase during routine use. A prospective registry-based observational cohort study from the United States reported nominal trends towards favorable clinical outcomes with tenecteplase 0.25 mg/kg over alteplase 0.9 mg/kg in the minor stroke (median NIHSS 4) subgroup, same as reported in the entire sample.^23^ Given the similarity in baseline characteristics of the ORIGINAL minor stroke subgroup to populations reported in a Chinese stroke surveillance report,^24^ and in an analysis of the Third China National Stroke Registry,^6^ the findings of this post hoc analysis are broadly generalizable across the entire patient group. The Third China National Stroke Registry analysis showed that IVT with alteplase versus non-IVT improved functional outcomes without increasing the risk of sICH in patients with minor stroke (NIHSS ≤5). The rate of mRS 0−1 (>80%) reported in the alteplase group was similar to that observed with both tenecteplase and alteplase in the current ORIGINAL minor stroke post hoc analysis. Compared to non-IVT, favorable functional outcomes persisted up to 1 year among minor stroke patients who received IVT.^6^

Certain guidances about IVT use in patients with minor stroke remain unclear, including the definition of minor stroke and treatment recommendations. While guidelines do not define a specific NIHSS cutoff for minor stroke, clinical trials often use NIHSS ≤5 although this threshold can vary.^25,26^ Moreover, the distinction between non-disabling and disabling minor stroke is based on clinical judgement regarding the functional impact of stroke.^10^ A disability perceived at presentation may or may not correspond with measurable deficit on the NIHSS or with future outcomes.^25,26^ A recent prospective cohort study leveraged data from the Third China National Stroke Registry to explore which patients with minor stroke might benefit most from alteplase. In a combined analyses of specific subgroups (NIHSS >3, disabling minor strokes, large artery atherosclerosis), IV alteplase was associated with a higher proportion of patients with excellent functional outcomes with no difference in in-hospital bleeding or mortality rates relative to standard medical treatment.^27^ Our post hoc analysis included patients with NIHSS scores ≤5, of whom those with scores <4 had motor dysfunction (disability). The results showed that these patients benefited from tenecteplase thrombolysis as effectively as the overall population.

Despite guideline recommendations, a relevant proportion of patients with minor disabling stroke are not treated with IV thrombolysis (e.g. 38% in a China National Stroke Registry analysis).^6^ possibly reflecting concerns about ICH. The ORIGINAL and AcT post hoc analyses both point to low rates of sICH with IVT in the minor stroke population.

Furthermore, guideline recommendations against IVT use in patients without clearly disabling symptoms are not wholly supported by the evidence. The PRISMS trial comparing intravenous alteplase with aspirin in patients with minor non-disabling deficits was terminated early by the sponsor due to slow patient enrolment, thus precluding any definitive conclusions.^28^ Patients with minor ischemic stroke and proven occlusion enrolled in the TEMPO-2 study were treated with tenecteplase or standard of care within 12 hours from stroke onset which exceeds the recommended time window for use of IV thrombolytics.^20^

Limitations of the ORIGINAL clinical trial have been reported elsewhere and include a lack of data on patients with large vessel occlusion and stroke mimics.^21^ As the post hoc analysis was not adequately powered to demonstrate noninferiority between the tenecteplase and alteplase treatment groups, the results must be interpreted with caution. This applies particularly to predefined subgroup analyses given the small sample sizes in certain subgroups. The results relate only to the comparability of tenecteplase and alteplase in minor disabling stroke since, to date, no RCTs have compared tenecteplase with standard treatment and antiplatelet therapy in this patient setting.

In conclusion, tenecteplase is endorsed by many groups as an alternative to alteplase for patients with AIS; and its logistical and pharmacological advantages suggest value with use in minor stroke. Aligning with results in the overall ORIGINAL population, this post hoc analysis indicated that the efficacy and safety of tenecteplase 0.25 mg/kg is comparable to that of alteplase 0.9 mg/kg in the minor stroke subpopulation (NIHSS ≤5), suggesting that tenecteplase is a suitable alternative to alteplase in this setting.

## Data Availability

Data Sharing Statement: To ensure independent interpretation of clinical study results and enable authors to fulfil their role and obligations under the International Committee of Medical Journal Editors (ICMJE) criteria, Boehringer Ingelheim grants all external authors access to relevant material, including participant-level clinical study data. In adherence with Boehringer Ingelheim Policy on Transparency and Publication of Clinical Study Data (see https://www.mystudywindow.com/msw/transparencypolicy), scientific and medical researchers can request access to clinical study data after publication of the primary manuscript in a peer-reviewed journal, providing regulatory activities are complete and other criteria met. Researchers should use the https://vivli.org/ link to request access to study data and visit https://www.mystudywindow.com/ for further information.

## Author Contributions

Shuya Li and Yongjun Wang contributed to the conception, design, and critical review of the article. Shuhong Xu drafted and revised the manuscript. Hongguo Dai, Guozhi Lu, Weiwei Wang, Fengyuan Che, Yu Geng, and Xiaolong Bao contributed to the overall review and revision of the manuscript. Shijia Yan was responsible for the statistical analysis. All authors approved the final manuscript and agreed to be accountable for all aspects of the work.

## Meeting Presentation

This post hoc analysis was presented at the 11th European Stroke Organisation Conference, 21−23 May 2025, Helsinki, Finland.

## Data Sharing Statement

To ensure independent interpretation of clinical study results and enable authors to fulfil their role and obligations under the International Committee of Medical Journal Editors (ICMJE) criteria, Boehringer Ingelheim grants all external authors access to relevant material, including participant-level clinical study data. In adherence with Boehringer Ingelheim Policy on Transparency and Publication of Clinical Study Data (see https://www.mystudywindow.com/msw/transparencypolicy), scientific and medical researchers can request access to clinical study data after publication of the primary manuscript in a peer-reviewed journal, providing regulatory activities are complete and other criteria met. Researchers should use the https://vivli.org/ link to request access to study data and visit https://www.mystudywindow.com/ for further information.

## Additional Contributions

Editorial assistance was provided by Kerry Dechant, ISMPP CMPP™, on behalf of Content Ed Net (Shanghai, China), and was funded by Boehringer Ingelheim China in accordance with Good Publication Practice (GPP2022) guidelines.

